# Estimation of the probability of reinfection with COVID-19 coronavirus by the SEIRUS model

**DOI:** 10.1101/2020.04.02.20050930

**Authors:** Victor Alexander Okhuese

**Affiliations:** Department of Mathematics, Nasarawa State University, Keffi – Nigeria

**Keywords:** coronavirus pandemic globally, coronavirus 2019-nCoV, mathematical modeling of infection disease, SEIRUS-model, parameter identification, statistical methods, COVID-19

## Abstract

With sensitivity of the Polymerase Chain Reaction (PCR) test used to detect the presence of the virus in the human host, the global health community has been able to record a great number of recovered population. Therefore, in a bid to answer a burning question of reinfection in the recovered class, the model equations which exhibits the disease-free equilibrium (***E*_0_**) state for COVID-19 coronavirus was developed in this study and was discovered to both exist as well as satisfy the criteria for a locally or globally asymptotic stability with a basic reproductive number *R*_0_ = 0 for and endemic situation. Hence, there is a chance of no secondary reinfections from the recovered population as the rate of incidence of the recovered population vanishes, that is, *B* = 0.

Furthermore, numerical simulations were carried to complement the analytical results in investigating the effect of the implementation of quarantine and observatory procedures has on the projection of the further spread of the virus globally. Result shows that the proportion of infected population in the absence of curative vaccination will continue to grow globally meanwhile the recovery rate will continue slowly which therefore means that the ratio of infection to recovery rate will determine the death rate that is recorded globally and most significant for this study is the rate of reinfection by the recovered population which will decline to zero over time as the virus is cleared clinically from the system of the recovered class.

## 1. INTRODUCTION

The COVID-19 coronavirus pandemic may have hit the world in a large scale as predicted in Victor (2020) and Batista (2020), the ratio of the death to recovery rate has seemingly been a positive proportion. However, with the sensitivity of the PCR, the presence or the absence of the virus in a previously infected host is detectable and as the recovery rate has seem to be encouraging considering the absence of any curative vaccine, the question in every quarters to health workers, the Center for Disease Control and the World Health Organization has been if reinfection could occur after a COVID-19 patient has recovered clinically?

A recent study by Victor (2020), Nesteruk (2020) and Ming and Zhang (2020) focuses on the epidemic outbreak cased by COVID-19 coronavirus due to the global trend of the pandemic with its origin from mainland China. In his study, Nesteruk (2020) used the popular SIR (Susceptible-Infectious-Removed) model to obtain optimal values for the model parameters with the use of statistical approach and hence predicated the number of infected, susceptible and removed persons versus time. This model approach by Nesteruk (2020) has been a major breakthrough in modelling disease control as used by several authors (Ming and Zhang, (2020) and Victor (2020) among others). However, although there exist a global interest in knowing the rate of infection that will occur over time globally, in this study we adopt solutions from Victor (2020) and Victor and Oduwole (2020) for a new deterministic endemic model (Susceptible – Exposed – Infectious – Removed – Undetectable – Susceptible: SEIRUS) originally developed for the control of the prevalence of HIV/AIDS in Africa.

The resulting equations are a system of coupled homogenous differential equations for projecting the rate of detection of the presence of the virus in the clinically prescribed recovered population. Numerical experiments with relevant simulation showing how the variation of the reproductive number (R_0_) affect the number of infected individuals is carried out as well as a projection for the rate of reinfection by the recovered class. Conscious effort through evaluating the new deterministic SEIRUS model is done to reduce the reproductive number (R_0_) to zero for a possible halt of the spread of the disease thereby leading to an endemic equilibrium to eradicate the disease in a later time in the future.

In summary, this study aims to use the new deterministic endemic SEIRUS compartmental model of the COVID-19 coronavirus dynamics which combines quarantine/observatory procedures and behavioral change/social distancing in the control and eradication of the disease in the most exposed sub-population to predict the chances of reinfection by the recovered class in the population.

The wide spread of the COVID-19 coronavirus and the lack or inefficiency of purposeful and result based intervention is a great call for other empirical and scientific interventions which seeks to review strategic models and recommendations of social and scientific research for disease control. Although previous studies have been tailored towards the epidemiology and the disease-free equilibrium where the reproductive number of the infectious population is at its barest minimum, this study seeks to study the evaluative impact of the endemic equilibrium of a new endemic deterministic model while taking into consideration that possibility of the recovered population being Undetectable and fit to be moved to the Susceptible compartment which will therefore imply a zero secondary infection of the disease globally.

Also, this research work will contribute significantly to recent research findings by Victor (2020), Nesteruk (2020) and Ming and Zhang (2020) as well as inform government, non-government organizations as well as policy maker’s decision on sustainable policies to prevent the spread of the disease based on specific age groups of the active population.

## 2. THE MODEL VARIABLES AND PARAMETERS

As suggested in Victor (2020), the model variables and parameters for the investigation of the stability analysis of the equilibrium state for the new deterministic endemic model is given by;

| Variable | Description |
| --- | --- |
| $S(t)$ | Number of susceptible population at time $t$ |
| $E(t)$ | Number of exposed population at time $t$ |
| $I(t)$ | Number of infected population at time $t$ |
| $R(t)$ | Number of infected population quarantined and expecting recovery at time $t$ |
| $U(t)$ | Number of recovered adults satisfying undetectable criteria at time $t$ |

| Parameter | Description |
| --- | --- |
| $\mu$ | Natural death rate of the population |
| $\alpha_0$ | Maximum death rate due to COVID-19 coronavirus. ($\alpha \leq \alpha_0$) |
| $\alpha$ | Death rate of infected population due to COVID-19 coronavirus |
| $\varphi$ | Disease induced death rate of infected population not quarantined |
| $\varpi$ | Disease induced death rate of infected receiving quarantined |
| $T$ | Maximum lifespan after infection ($T \geq 14 \text{ days}$) |
| $k$ | Efficacy of Quarantine ($0 \leq k \leq 1$) |
| $\rho$ | Rate of recovery |
| $\beta$ | rate of transmission |
| $\sigma$ | Proportion of infected population in quarantine per unit time (Treatment rate) |
| $\pi$ | Proportion of population from susceptible to exposed/latent class |
| $\varepsilon$ | Proportion of removed population still being observed and being moved to susceptible class |
| $B(t)$ | Incidence rate or force of infection in the population |

### 2.1 MODEL ASSUMPTIONS

The following assumptions as suggested in Victor (2020) and Victor and Oduwole (2020) helps in the derivation of the model:

1. There is no emigration from the total population and there is no immigration into the population. A negligible proportion of individuals move in and out of the population at a given time.
2. Maturation (or maturity) is interpreted as the period between infection to the period of symptoms observation (days 1 to 14)
3. The susceptible population are first exposed to a latent class where they can infected or not.
4. Some infected individuals move to the removed class when they are quarantined for observatory procedures.
5. The recruitment from the *S*-class into the *E*-class is through contacts from population in the *I*-class to the *S-*class
6. The recruitment into the *R*-class from the *I*-class at a rate *σ*.
7. The recruitment into the *U*-class from the *R*-class depends on the effectiveness of the quarantine and observatory procedure at a rate *ρ*.
8. Death is implicit in the model and it occurs in all classes at constant rate *μ*. However, there is an additional death rate in the *I* and *R* classes due to infection for both juvenile and adult sub-population denoted by *φ* and *ϖ* respectively.

### 2.2 MODEL DESCRIPTION

This study uses the deterministic endemic model where a susceptible class is a class that is yet to be infected, but is open to infection as interactions with members of the *I*-class continuous. An infected individual is one who has contracted the coronavirus and is at some stage of infection. A removed individual is one that is confirmed to have the virus with its expected symptoms and is under quarantine while following relevant observatory procedures. A member of the undetectable class is one that has been removed and does not secrete the virus anymore and has been satisfied by the WHO standard to be in the undetectable class.

The following diagram describes the dynamic of SEIRUS framework, and will be useful in the formulation of model equations.

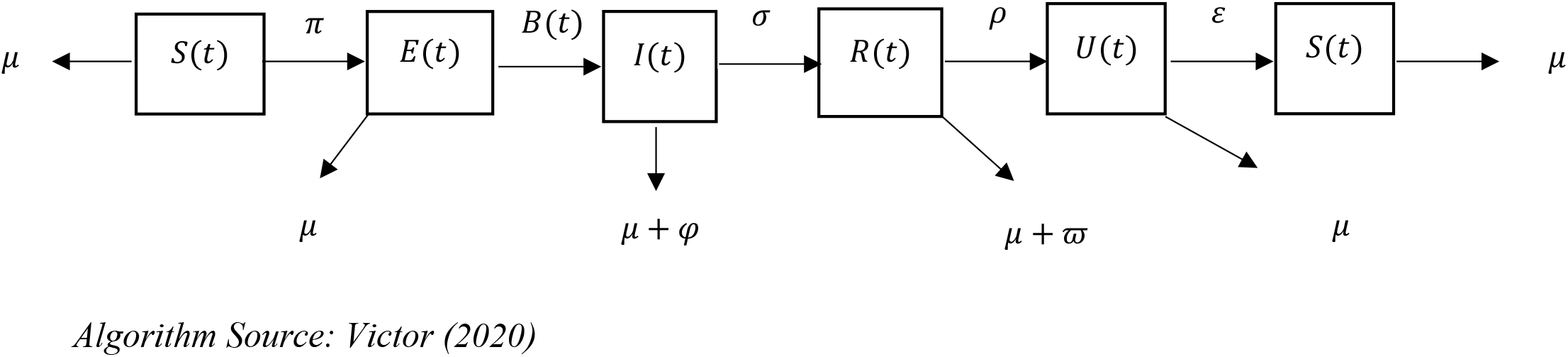

## 3. THE MODEL EQUATIONS

The resulting equations below are a system of coupled homogenous differential equations for projecting the rate of detection of the presence of the virus in the clinically prescribed recovered population based on the assumptions and the flow diagram above.

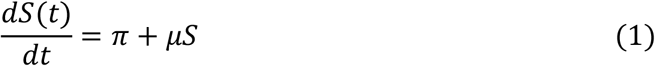

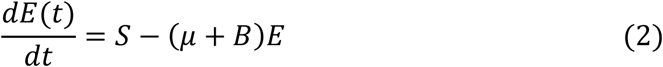

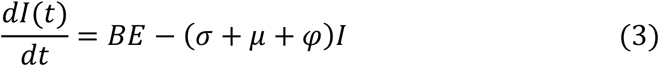

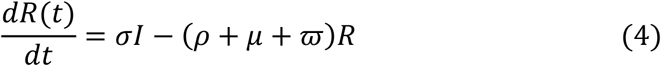

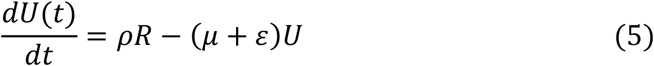

Such that

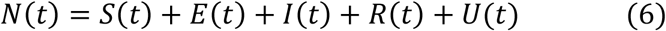

The incidence rate or force of infection at time *t* denoted by *B*(*t*) in the population is given as

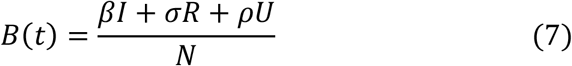

### 3.1 MODEL EQUATIONS IN PROPORTIONS

The model equations in proportion according to Victor (2020) is adopted for this study as follows;

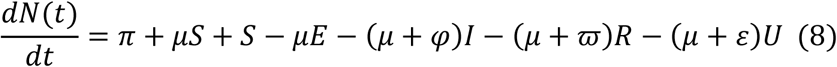

Let

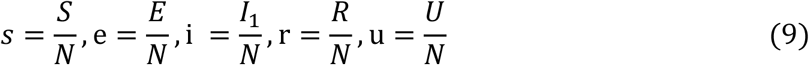

Such that;

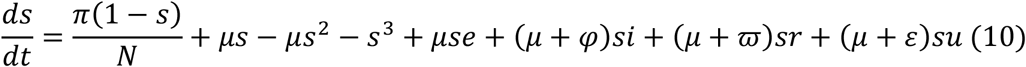

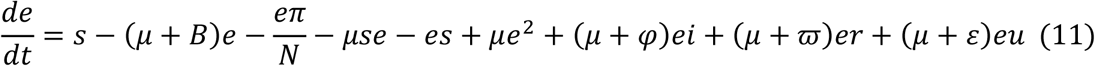

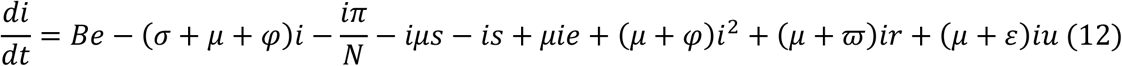

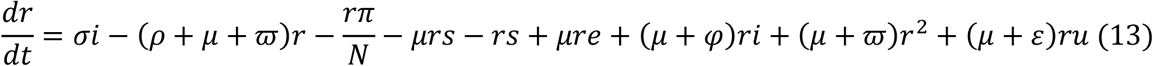

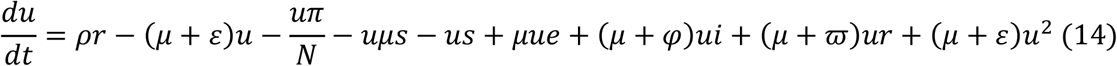

However,

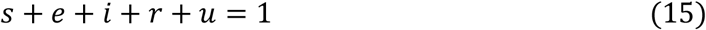

Equations (10) to (14) are the model equations in proportions, which define prevalence of infection.

### 3.2 EXISTENCE AND UNIQUENESS OF DISEASE FREE EQUILIBRIUM STATE (*E*_0_) OF THE SEIRUS MODEL

The disease-free equilibrium (DFE) state of the endemic SEIRUS model is obtained by setting the left hand sides of equations (10) – (14) to zero while setting the disease components *e* = *i* = *r* = *u* = 0 leading to equations (15) – (16) below

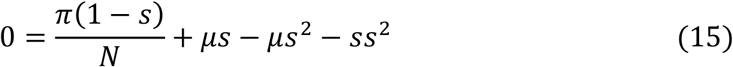

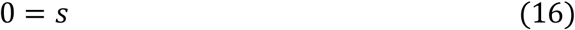

And substituting (16) into (15) we have;

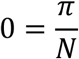

Which makes

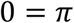

Then taking (15) where *s* = 0 or

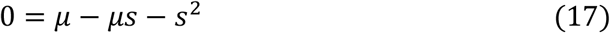

Simplifying further gives,

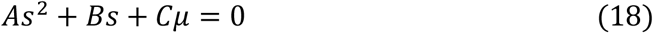

where *A* = *1, B* = *μ* and *C* = *−μ*

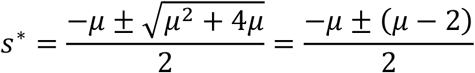

Therefore, the solution for the simultaneous equations (18) is given by

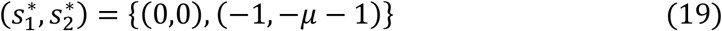

Ignoring the native values of 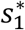 and 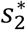 and other stringent conditions, there exist a unique trivial and disease-free equilibrium states at 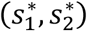 given by (0,0). The solution (19) satisfies equation (18) identically.

### 3.3 STABILITY ANALYSIS OF DISEASE FREE EQUILIBRIUM STATE (*E*_0_) RECOVERED POPULATION

In the event where patients recover from the COVID-19 coronavirus, it is assumed so far that they are disease-free at least from 14 days after their last clinical test shows that they have clinically recovered from the virus. Hence, in order to study the behavior of the systems (10) – (14) around the disease-free equilibrium state *E*_0_ = [0,0,0,0,0], we resort to the linearized stability approach from Victor (2020) gives us a Jacobian 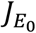 transformation of the form;

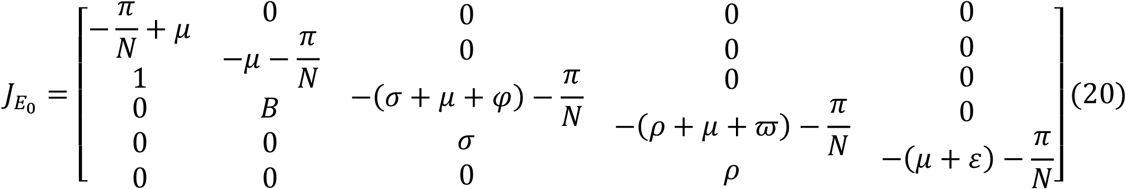

Hence, according to (Gerald, 2012, p255), the Determinant of the Jacobian Matrix 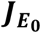 is given by the recursive definition for a 5 × 5 matrix defined as;

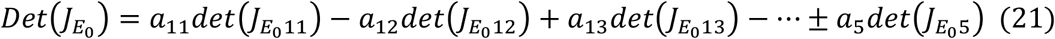

And from (20)

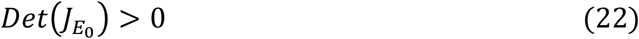

Similarly from the Trace of the Jacobian matrix (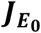) given in equation (20) we have that

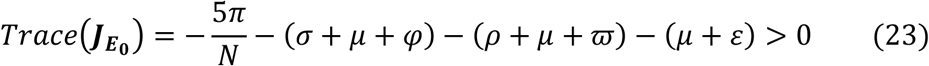

Hence, since 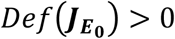 and 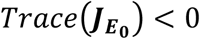 which does satisfy the prescribed threshold criteria based on Gerald (2012), then the disease free equilibrium (***E*_0_**) for COVID-19 coronavirus does satisfy the criteria for a locally or globally asymptotic stability for the recovered population. This implies that as a pandemic as declared by WHO (2020) the COVID-19 coronavirus does not have a curative vaccine yet and precautionary measures are advised through quarantine and observatory procedures. Therefore, for the recovered population the chances reinfection appears to be uncertain though closely impossible unless regular clinical test is not accurately administered.

### 3.4 COMPUTATION OF THE BASIC REPRODUCTIVE NUMBER (*R*_0_) OF THE MODEL

The Basic Reproductive number (*R*_0_) is define as the number of secondary infections that one infectious individual would create over the duration of the infectious period, provided that everyone else is susceptible. *R*_0_ = *1* is a threshold below which the generation of secondary cases is insufficient to maintain the infection in human community. If *R*_0_ *< 1*, the number of infected individuals will decrease from generation to next and the disease dies out and if *R*_0_ *> 1* the number of infected individuals will increase from generation to the next and the disease will persist. To compute the basic reproductive number (*R*_0_) of the model with incidence rate for the recovered population assumed to vanish such that *B* = 0 and we employ the next generation method as applied by Diekmann *et. al*., (2009), Van den Driessche and Watmough (2002).

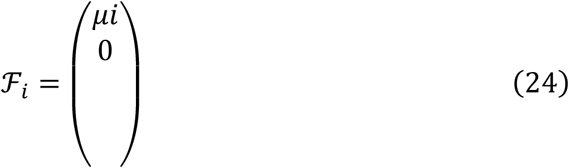

and

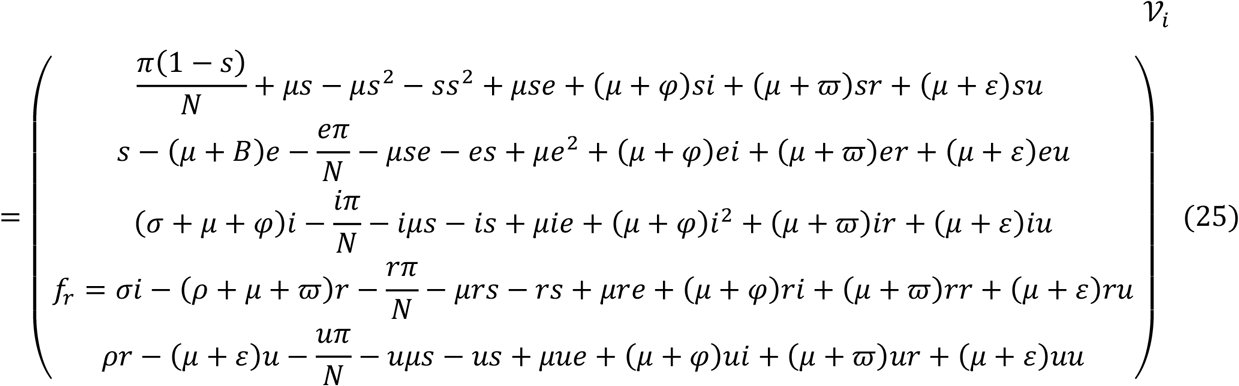

where ℱ_*i*_ and *𝒱*_*i*_ are the rate of appearances of new infections in compartment *i* and the transfer of individuals into and out of compartment *i* by all means respectively. Using the linearization method, the associated matrices at disease-fee equilibrium (*E*_0_) and after taking partial derivatives as defined by

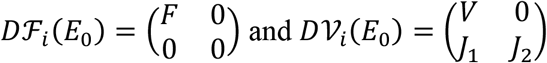

where *F* is nonnegative and *V* is a non-singular matrix, in which both are the *m × m* matrices defined by

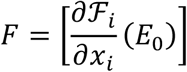

And

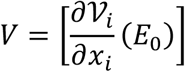

with *1 ≤ i, j ≤ m* and *m* is the number of infected classes.

In particular *m* = 2, we have

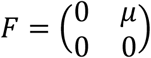

and

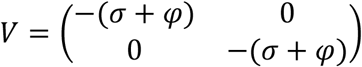

If the inverse of *V* is given as

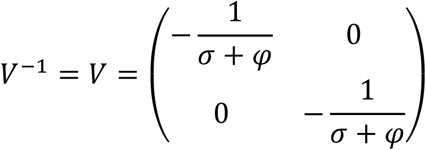

Then the next matrix denoted by *FV*^*−1*^ is given as

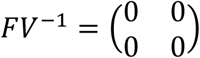

We find the eigenvalues of *FV*^*−1*^ by setting the determinant |*FV*^*−1*^ *− γI*| = 0

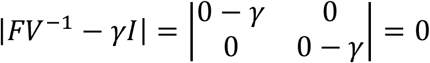

with characteristics polynomial

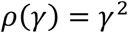

and characteristics equation given as

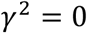

Solving the characteristics equation for the eigenvalues *γ*_1,2_, where *R*_0_ is the maximum of the two eigenvalues *γ*_*1,2*_. Hence the Basic Reproductive number is the dominant eigenvalues of *FV*^*−1*^. Thus we have that

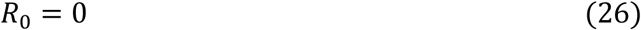

The Basic Reproductive number (*R*_0_ = 0) by Equation (33) shows that with no incidence rate in the recovered population, there is no chances secondary infection by COVID-19 coronavirus patients who have been clinically declared negative and free from the virus with the virus completely cleared from their system. Hence although there currently exist no clinical vaccine for the cure of COVID-19 coronavirus, with Equation (33) there is a high chance of no case of reinfection after clinical recovery from the virus.

## 4. DESCRIPTION AND VALIDATION OF BASELINE PARAMETERS FOR GLOBAL CASES OF COVID-19 CORONAVIRUS

According to the WHO (2020), the total cases of COVID-19 coronavirus worldwide stands: about 900,000, with a total of about 190,000 recovery and the current total deaths is about 44,000 from about 172 countries.

### 4.1 NUMERICAL EXPERIMENTS OF THE MODEL

The age-structured deterministic model (10) – (14) was solved numerical using Runge-Kutta-Fehllberg 4-5th order method and implemented using Maple Software. The model equations were first transformed into proportions, thus reducing the model equations to ten differential equations. The parameters used in the implementation of the model are shown in Table 1 below. Parameters were chosen in consonance with the threshold values obtained in the stability analysis of the disease free equilibrium state of the model.

**Table 1:**
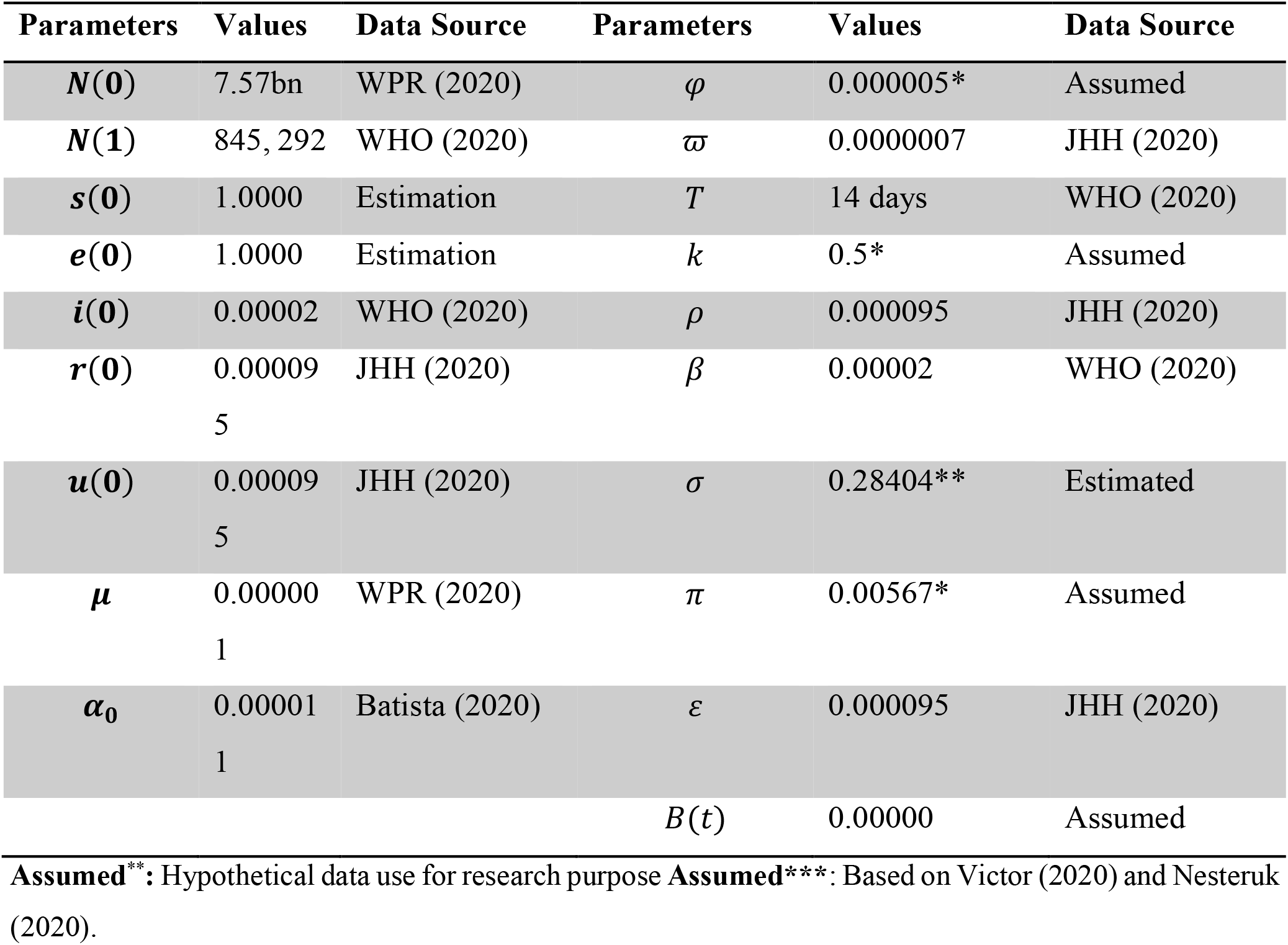
Estimated values of the parameters used in the Numerical experiments

Hence from Equation (26) the Reproductive Number *R*_0_ = 0 means there is a 100% chances of zero secondary reinfection from the recovered compartment of the COVID-19 patient group when a re-infectious population interact by contact with the susceptible population.

**Figure 1:**
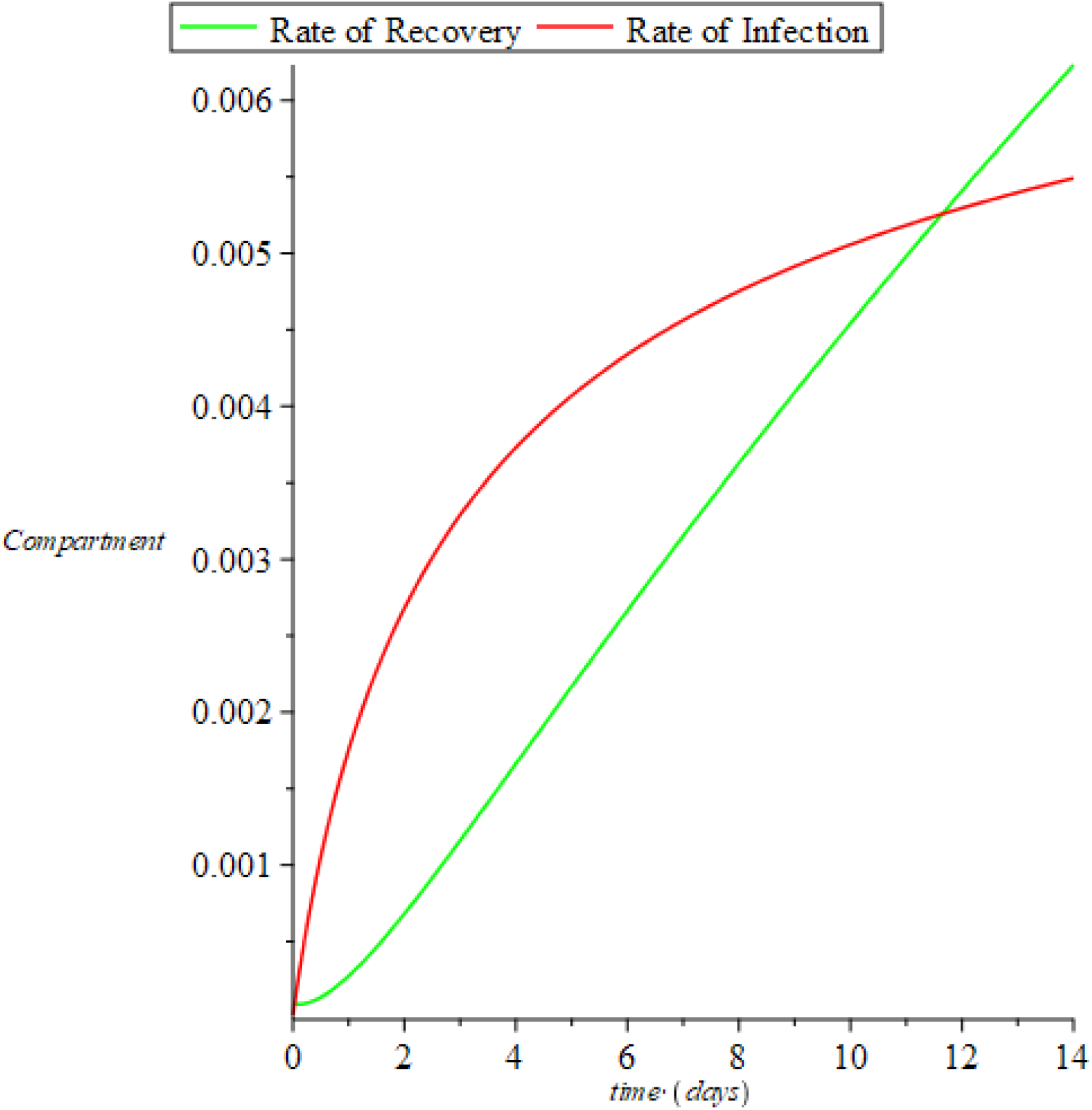
Chart of Recovered and Infectious compartment for the COVID-19.

**Figure 2:**
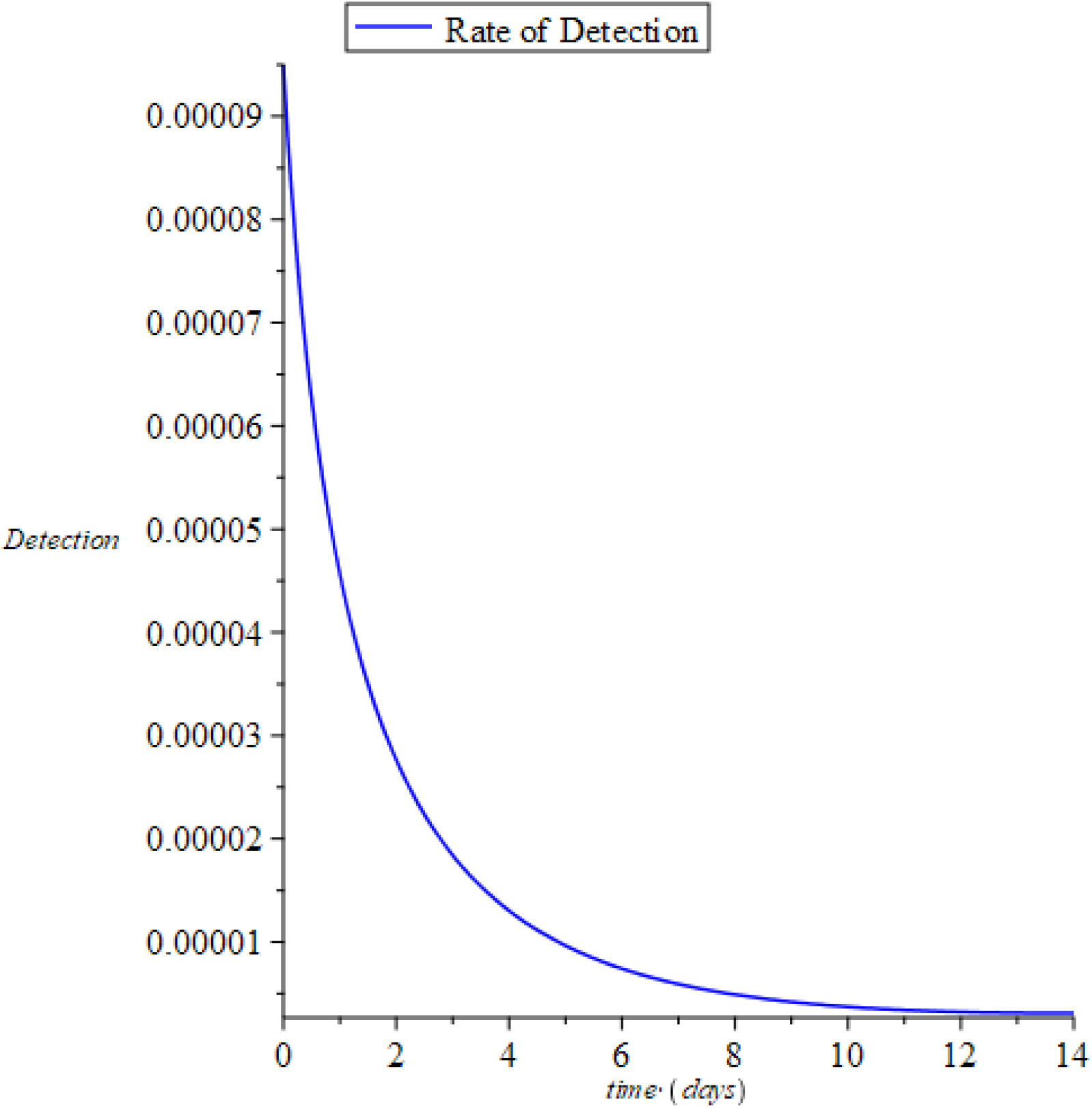
Chart of the rate of reinfection of the recovered compartment from COVID-19

Figure 1, clearly shows that the infectious rate will continue to rise steadily over a long period of time and there after begin to slide in a normal trajectory if no vaccine is in place. However, with the various make shift treatment, social distancing and quarantine strategy being adopted the recovery rate will keep rising slowly but steadily over a long period of time. Therefore, as the recovery rate continues to grow steadily the number of recovered population who have been clinically declared free of the virus by the Polymerase Chain Reaction test will be also declared un-infectious as long as the virus is completely cleared from their system (see Figure 2) and the rate of detection will vanish, making the rate of secondary infection *R*_0_ = 0 as long as the incidence rate *B* = 0.

**Figure 1:**
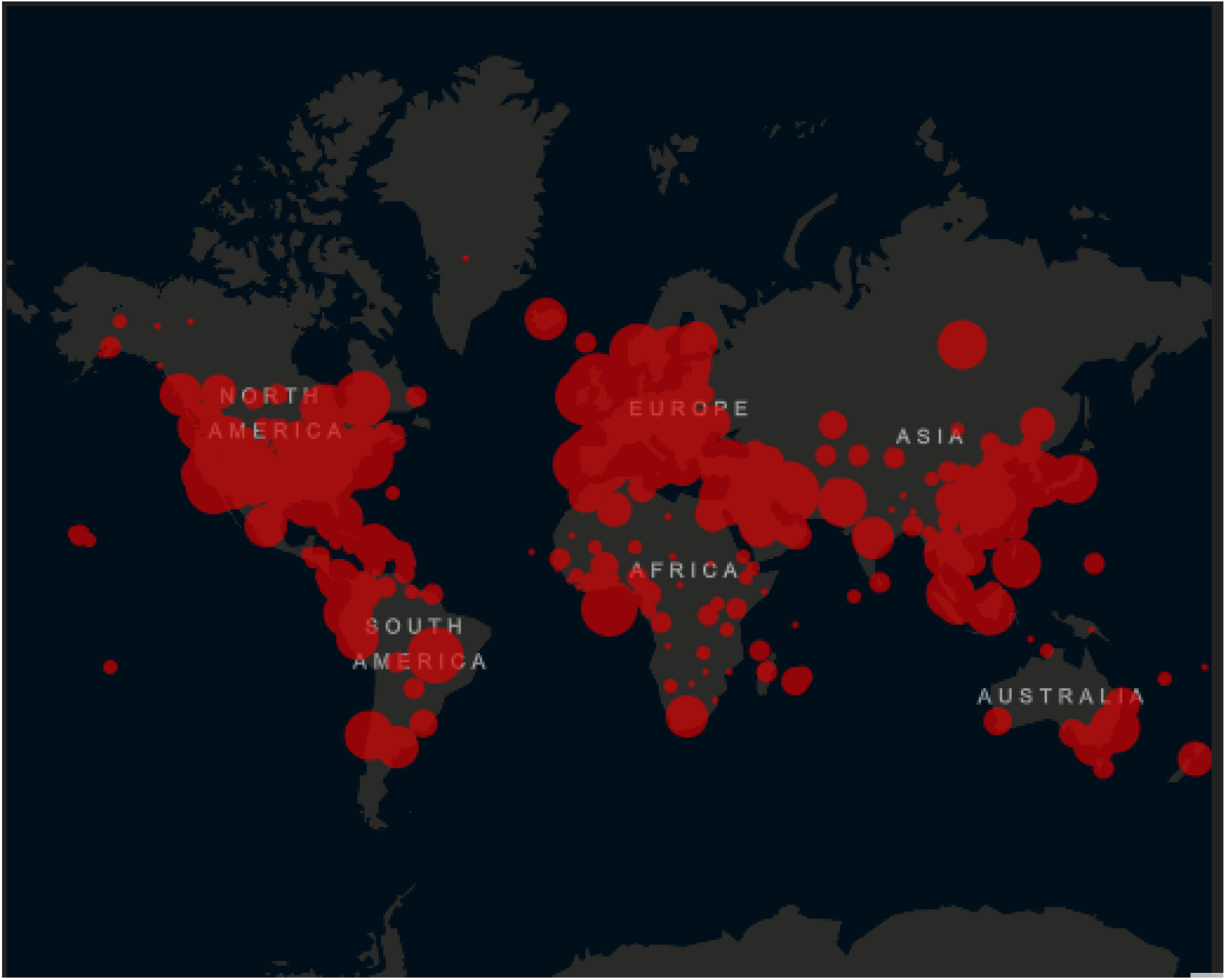
A World Map showing the number of cases per each country with a COVID-19 coronavirus case. (*Source: JHU (2020)*).

**Figure 2:**
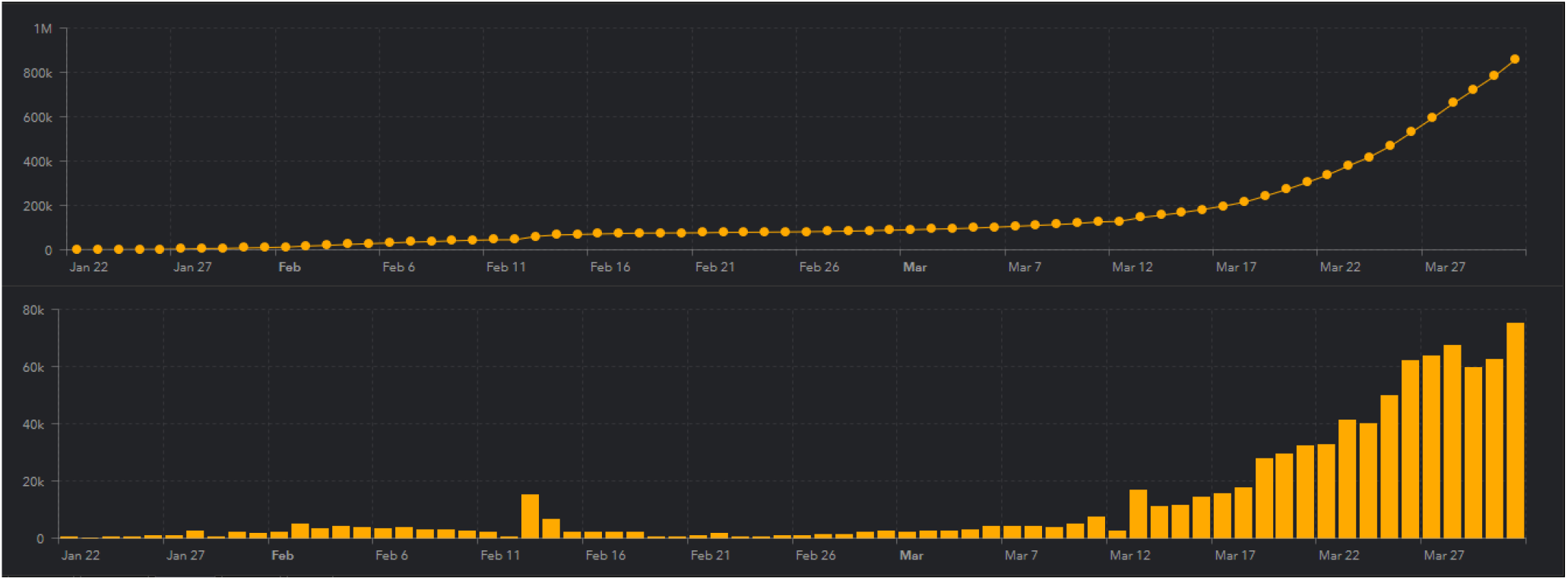
A Cumulative Case chart showing the number of cases with a COVID-19 coronavirus case. (*Source: JHU (2020)*).

## CONCLUSIONS

The model equations which exhibits the disease-free equilibrium (***E*_0_**) state for COVID-19 coronavirus exist and hence satisfies the criteria for a locally or globally asymptotic stability when the basic reproductive number *R*_0_ = 0 for and endemic situation. This implies that as a pandemic as declared by WHO (2020) the COVID-19 coronavirus does not have a curative vaccine yet and precautionary measures are advised through quarantine and observatory procedures. The basic reproductive number was found to be *R*_0_ = 0 and hence shows that there is a high chance of no secondary infections from the recovered population as the rate of incidence of the recovered population vanishes, that is, *B* = 0.

However, unless there is a dedicated effort from individual population, government, health organizations, policy makers and stakeholders, the world would hardly be reed of the COVID-19 coronavirus and further spread is eminent and the rate of infection will continue to increase despite the increased rate of recovery until a curative vaccine is developed.

## Data Availability

Data available for this study were estimated by the author and others were retrieved from the WPR (2020), WHO (2020) and JHU (2020) which were publicly available data.

